# Smell, taste and chemesthesis disorders in patients with the SARS-CoV-2 Omicron variant in China

**DOI:** 10.1101/2023.02.21.23286242

**Authors:** Ying Chen, Yuying Chen, Xiang Liu, Chao Yan, Laiquan Zou

**Affiliations:** Chemical Senses and Mental Health Lab, Department of Psychology, School of Public Health, Southern Medical University, Guangzhou, Guangdong, China; Department of Otolaryngology, Sun Yat-sen Memorial Hospital, Sun Yat-sen University, Guangzhou, Guangdong, China; Key Laboratory of Brain Functional Genomics (MOE & STCSM), Shanghai Changning-ECNU Mental Health Center, School of Psychology and Cognitive Science, East China Normal University, Shanghai, China

**Keywords:** chemosensory, COVID-19, cross-sectional studies, Omicron, prevalence

## Abstract

**Background:** Chemosensory disorders (including smell, taste and chemesthesis) are among the established symptoms of COVID-19 infection; however, new data indicate that the changes in chemosensory sensation caused by COVID-19 may differ among populations and COVID-19 variants. To date, few studies have focused on the influence of the SARS-CoV-2 Omicron variant on qualitative changes and quantitative reductions in chemosensory function in China.

**Methodology:** We conducted a cross sectional study of patients with COVID-19 caused by the Omicron variant, to investigate the prevalence of chemosensory disorders and chemosensory function before and during infection, using an online questionnaire.

**Results:** A total of 1245 patients with COVID-19 completed the survey. The prevalence rates of smell, taste, and chemesthesis disorders were 69.2%, 67.7%, and 31.4%, respectively. Our data indicate that sex, age, smoking, and COVID-19-related symptoms, such as lack of appetite, dyspnea, and fatigue, may be associated with chemosensory disorders during COVID-19.

**Conclusions:** Self-rating of chemosensory function revealed that patients experienced a general decline in smell, taste, and chemesthesis function. Further longitudinal research studies are needed to generate additional data based on objective assessment and investigate the factors influencing chemosensory function in COVID-19.

## INTRODUCTION

Coronavirus disease 2019 (COVID-19) was first detected in Wuhan in December 2019, and rapidly spread throughout more than 80 countries ^(1,2)^. Information from the World Health Organization shows that the number of confirmed COVID-19 cases was 756,411,740 by 16 February, 2023 (https://covid19.who.int). There are reports that chemosensory disorders are important symptoms of COVID-19 infection ^(3–5)^. Studies from the United States, Europe, Malaysia, and Singapore demonstrated that 12%– 88% of COVID-19-infected patients have olfactory and taste disorders ^(4,6–8)^, while studies from China reported that 5%–25% of patients have smell and/or taste disorder ^(9,10)^. The Omicron variant of SARS-CoV-2 was first detected in South Africa in November 2021, and became the main epidemic strain in the world by 15 January 2022 ^(11,12)^. Some studies have found that the Omicron variant causes less smell and taste dysfunction than non-Omicron SARS-CoV-2 ^(13–15)^. Further, a meta-analysis found that the prevalence rates of olfactory dysfunction caused by the Omicron variant are 8%–17% and 2%–17% the in UK and USA, respectively ^(15)^.

In China, the prevalence of olfactory dysfunction caused by the Omicron variant is reported as 0–9% ^(16–20)^; however, the sample sizes of studies to date have been relatively small and researchers have focused on patient characteristics and clinical symptoms, with insufficient discussion of chemosensory changes.

Chemosensory sensation (including smell, taste, and chemesthesis) have important roles in sensing potential threats, such as toxins, bacteria, and chemical irritants ^(21)^, but most relevant studies have focused solely on smell and taste, and ignored chemesthesis, which is defined as detection of burning, cooling, or tingling in the mouth ^(5,21)^. Since these three systems are independent sensory systems, with distinct peripheral and central neural mechanisms ^(22)^, it is necessary to explore the influence of COVID-19 on these three senses separately. Further, chemosensory disturbances can result in qualitative changes or quantitative reductions in smell or taste, associated with different mechanisms. Qualitative changes include parosmia (things smell different from usual), phantosmia (hallucination of olfactory senses), smell fluctuation, parageusia (distorted taste sensations) and phantogeusia (hallucination of gustatory senses). Quantitative reductions include anosmia (complete loss of olfaction), hyposmia (partial loss of olfaction), ageusia (loss of all or specific gustatory senses) and hypogeusia (reduced ability to taste things) ^(23,24)^. Previous studies have generally explored changes or reductions of smell and taste, but have rarely classified patient conditions according to the qualitative of the disorders.

There is a pressing need to conduct a cross-sectional study to assess the effects of Omicron on patient smell, taste, and chemesthesis in China. The primary aim of this study was to investigate the prevalence of chemosensory disorders, as well as qualitative and quantitative changes of the chemosensory senses during Omicron infection. A secondary aim was to identify factors associated with chemosensory disorders, including sex, age, alcohol intake, chronic rhinitis, allergic rhinitis, and comorbidity or specific conditions, as well as COVID-19 related characteristics.

## MATERIALS AND METHODS

### Participants

Participants answered questions in an online survey about their sociodemographic characteristics, COVID-19 infection status, and smell, taste, and chemesthesis between 20 December 2022 and 20 January 2023. A total of 1311 patients (all aged ≥ 18 years) recovered from COVID-19 were invited to participate in the survey. Participants who did not answer all of the questions or failed to pass the lie test were excluded. The final sample consisted of 1245 participants: 983 women (79.0%) and 262 men (21.0%), with a mean ± standard deviation (SD) age of 25.45 ± 6.57 years old. As this was a cross-sectional study, a 0.95 power estimate, an effect size of 0.3, and an α value of 0.05 were used to calculate the necessary sample size in the G*Power program ^(25)^. The sample size in our study was far larger than the result proposed by the G*Power program. All participants read an informed consent form and agreed to the use of their data for research. This study received ethics approval from the Ethics Committee of Southern Medical University.

### Data collection

A self-administered questionnaire was used to survey patients who had recovered from COVID-19 in China. The questionnaire was adapted from existing online questionnaires developed by the Global Consortium for Chemosensory Research (GCCR) ^(5)^. The GCCR core questionnaire has been implemented in 10 languages as of April 18, 2020, and used or adapted in many other research studies ^(4,26,27)^. Data collected included demographic information, COVID-19-related characteristics, and patient chemosensory situations, before and during infection. To study the sense of smell and taste in further detail, smell situation options included anosmia, hyposmia, parosmia, phantosmia, and smell fluctuation, and taste situation options included ageusia, hypogeusia, parageusia, and phantogeusia ^(5,24)^.

### Statistical analysis

Associations between categorical variables were tested using the Chi-Square test. Self-ratings of smell, taste, and chemesthesis before and during COVID-19 diagnosis were evaluated using the Wilcoxon matched pairs signed-rank test. Statistical significance was set at p < 0.05 and all reported p values are two-tailed. All statistical analyses were performed using IBM SPSS Statistics (SPSS, version 22).

## RESULTS

### General prevalence

Of 1245 patients infected with COVID-19, 69.2% (861/1245) reported having smell disorder, 67.7% (843/1245) reported having taste disorder, and 31.4% (391/1245) reported chemesthesis disorder (Table 1); the prevalence rates of specific types of smell and taste disorder are also shown in Table 1. Among patients who experienced anosmia or hyposmia (n = 819), only 12.3% (101/819) reported that the anosmia or hyposmia was completely caused by nasal congestion. Further, 56.9% (708/1245) of patients reported loss of both smell and taste, 12.3% (153/1245) reported loss of smell only, and 10.8% (135/1245) reported loss of taste only.

**Table 1.** The prevalence of smell, taste, and chemesthesis disorders among patients with COVID-19.

| Variable | n | Total prevalence (N = 1245) | Prevalence in people with smell/taste/chemesthesis disorder |
| --- | --- | --- | --- |
| Smell disorder* | 861 | 69.2% | \ |
| Anosmia | 290 | 23.3% | 33.7% |
| Hyposmia | 529 | 42.5% | 61.4% |
| Parosmia | 60 | 4.8% | 7.0% |
| Phantosmia | 98 | 7.9% | 11.4% |
| Smell fluctuation | 438 | 35.2% | 50.9% |
| Taste disorder* | 843 | 67.7% | \ |
| Ageusia | 236 | 19.0% | 28.0% |
| Hypogeusia | 499 | 40.1% | 59.2% |
| Parageusia | 282 | 22.7% | 33.5% |
| Phantogeusia | 333 | 26.7% | 39.5% |
| Chemesthesis disorder | 391 | 31.4% | \ |
| Complete loss | 35 | 2.8% | 9.0% |
| Partial loss | 356 | 28.6% | 91.0% |

\*Multiple responses

### Smell disorder

The data presented in Table 2 show that there was a statistically significant difference in smoking status between patients with and without smell disorder (p < 0.05); where smell disorder was significantly more common among patients who were current or former smokers (p < 0.05). No statistically significant differences in other demographic characteristics, including sex, age group, alcohol intake, chronic rhinitis, allergic rhinitis, and comorbidity or special condition, were found between patients with and without smell disorder (all *p* > 0.05). Significant differences in rehabilitation status, COVID-19-related symptoms (lack of appetite, dry and sore throat, myalgia, stuffy/running nose, dyspnea and fatigue), and COVID-19 vaccination status, were also observed between patients with and without smell disorder (all *p* < 0.05). Chi-square analysis indicated that patients with symptoms including lack of appetite, dry and sore throat, myalgia, stuffy/running nose, dyspnea, and fatigue, were prone to having smell disorder. Further, patients who did not undergo COVID-19 vaccination were more likely to experience smell disorder (p < 0.05). Associations between specific types of smell disorder (anosmia, hyposmia, parosmia, phantosmia, and olfactory fluctuation) and demographic/COVID-19-related characteristics are presented in the supplemental material (Table S1–S5).

**Table 2.**
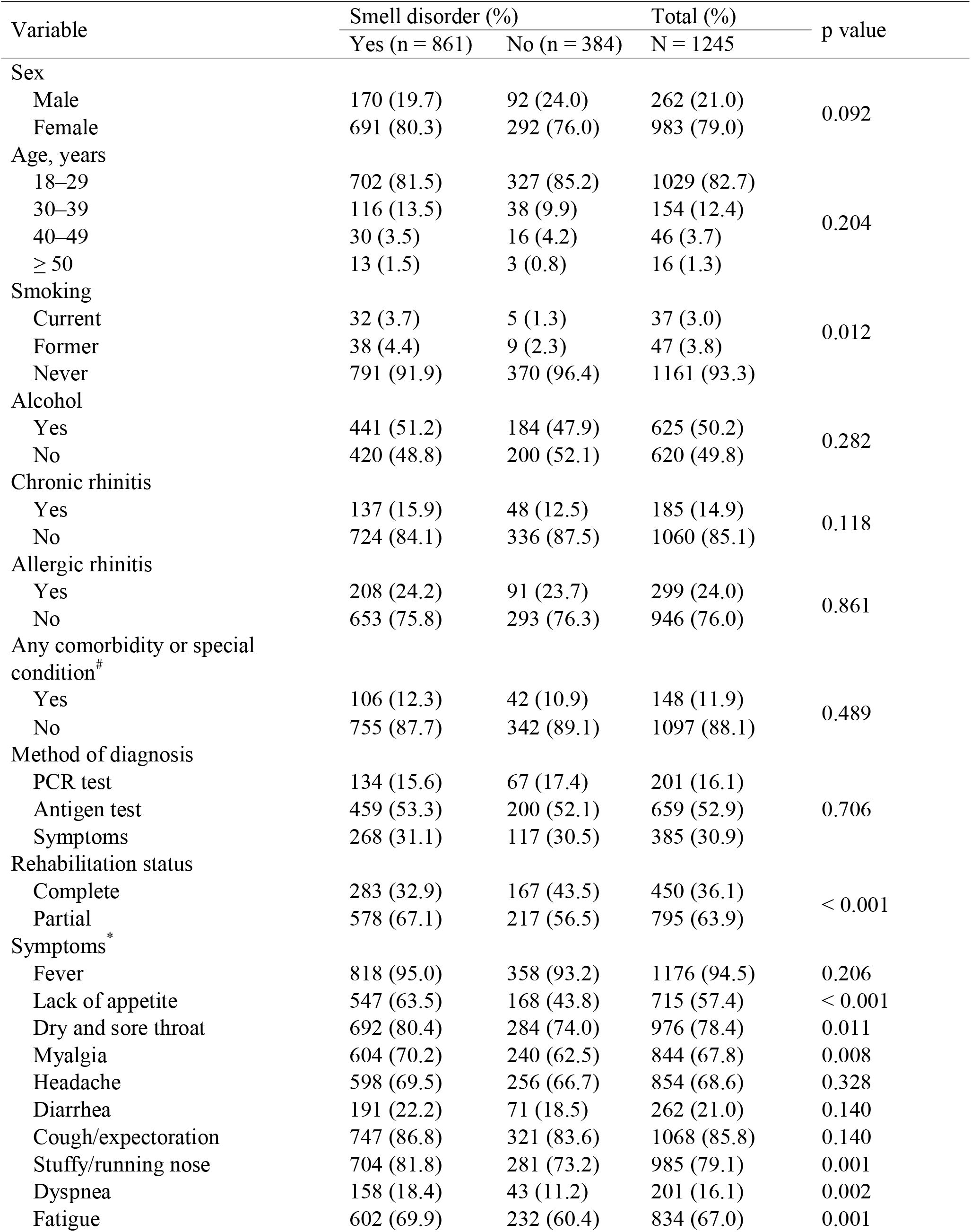
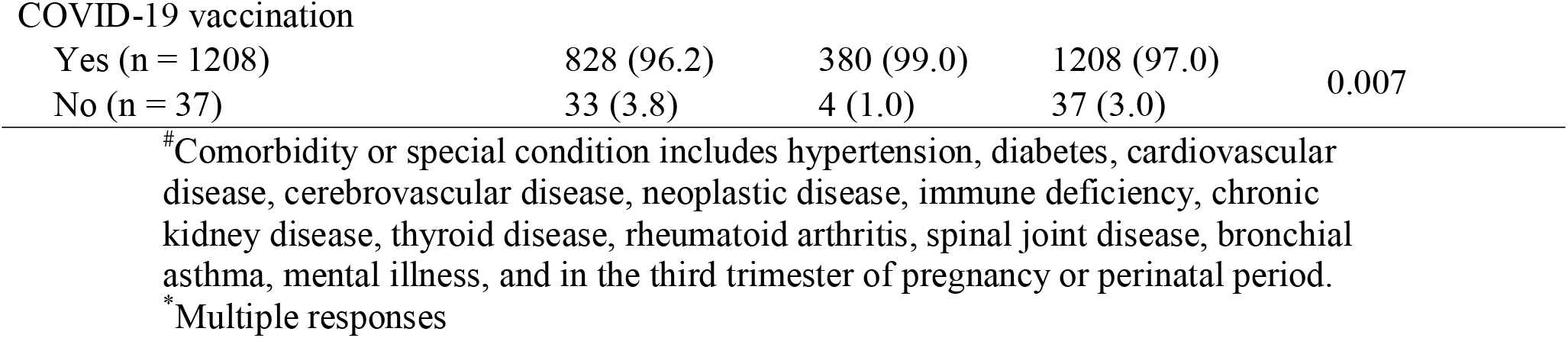
Association between smell disorder and demographic/COVID-19-related characteristics among patients.

| Variable | Smell disorder (%) |  | Total (%) | p value |
| --- | --- | --- | --- | --- |
|  | Yes (n = 861) | No (n = 384) | N = 1245 |  |
| Sex |  |  |  |  |
| Male | 170 (19.7) | 92 (24.0) | 262 (21.0) | 0.092 |
| Female | 691 (80.3) | 292 (76.0) | 983 (79.0) |  |
| Age, years |  |  |  |  |
| 18–29 | 702 (81.5) | 327 (85.2) | 1029 (82.7) | 0.204 |
| 30–39 | 116 (13.5) | 38 (9.9) | 154 (12.4) |  |
| 40–49 | 30 (3.5) | 16 (4.2) | 46 (3.7) |  |
| ≥ 50 | 13 (1.5) | 3 (0.8) | 16 (1.3) |  |
| Smoking |  |  |  |  |
| Current | 32 (3.7) | 5 (1.3) | 37 (3.0) | 0.012 |
| Former | 38 (4.4) | 9 (2.3) | 47 (3.8) |  |
| Never | 791 (91.9) | 370 (96.4) | 1161 (93.3) |  |
| Alcohol |  |  |  |  |
| Yes | 441 (51.2) | 184 (47.9) | 625 (50.2) | 0.282 |
| No | 420 (48.8) | 200 (52.1) | 620 (49.8) |  |
| Chronic rhinitis |  |  |  |  |
| Yes | 137 (15.9) | 48 (12.5) | 185 (14.9) | 0.118 |
| No | 724 (84.1) | 336 (87.5) | 1060 (85.1) |  |
| Allergic rhinitis |  |  |  |  |
| Yes | 208 (24.2) | 91 (23.7) | 299 (24.0) | 0.861 |
| No | 653 (75.8) | 293 (76.3) | 946 (76.0) |  |
| Any comorbidity or special condition<sup>#</sup> |  |  |  |  |
| Yes | 106 (12.3) | 42 (10.9) | 148 (11.9) | 0.489 |
| No | 755 (87.7) | 342 (89.1) | 1097 (88.1) |  |
| Method of diagnosis |  |  |  |  |
| PCR test | 134 (15.6) | 67 (17.4) | 201 (16.1) | 0.706 |
| Antigen test | 459 (53.3) | 200 (52.1) | 659 (52.9) |  |
| Symptoms | 268 (31.1) | 117 (30.5) | 385 (30.9) |  |
| Rehabilitation status |  |  |  |  |
| Complete | 283 (32.9) | 167 (43.5) | 450 (36.1) | < 0.001 |
| Partial | 578 (67.1) | 217 (56.5) | 795 (63.9) |  |
| Symptoms<sup>*</sup> |  |  |  |  |
| Fever | 818 (95.0) | 358 (93.2) | 1176 (94.5) | 0.206 |
| Lack of appetite | 547 (63.5) | 168 (43.8) | 715 (57.4) | < 0.001 |
| Dry and sore throat | 692 (80.4) | 284 (74.0) | 976 (78.4) | 0.011 |
| Myalgia | 604 (70.2) | 240 (62.5) | 844 (67.8) | 0.008 |
| Headache | 598 (69.5) | 256 (66.7) | 854 (68.6) | 0.328 |
| Diarrhea | 191 (22.2) | 71 (18.5) | 262 (21.0) | 0.140 |
| Cough/expectorations | 747 (86.8) | 321 (83.6) | 1068 (85.8) | 0.140 |
| Stuffy/running nose | 704 (81.8) | 281 (73.2) | 985 (79.1) | 0.001 |
| Dyspnea | 158 (18.4) | 43 (11.2) | 201 (16.1) | 0.002 |
| Fatigue | 602 (69.9) | 232 (60.4) | 834 (67.0) | 0.001 |
| COVID-19 vaccination |  |  |  |  |
| Yes (n = 1208) | 828 (96.2) | 380 (99.0) | 1208 (97.0) | 0.007 |
| No (n = 37) | 33 (3.8) | 4 (1.0) | 37 (3.0) |  |

Only 4.8% (60/1245) of patients reported parosmia, while 7.9% (98/1245) reported phantosmia. Pleasantness rating score on 100-point scale of the distorted smell caused by parosmia was 25.72 ± 24.81 (Mean ± SD), and that of phantom smells was 26.74 ± 25.98 (Mean ± SD). Parosmic individuals mostly reported smells of food (e.g., meat, rice, or soup) or other daily necessities (e.g., toothpaste, shampoo, or liquid detergent) that became roasted, burnt, or spoiled (e.g., “The smell of chicken soup turned to burning”). In phantosmic individuals, the most frequently reported phantom smells were smoky, burnt, and rotten.

Patients rated their smell function before and during COVID-19 on a 100-point scale. Compared to scores before COVID-19 (88.53 ± 13.24, Mean ± SD), those during COVID-19 (52.38 ± 32.83, Mean ± SD) were significantly lower (t = 38.386, p < 0.001) (Figure 1).

**Figure 1.**
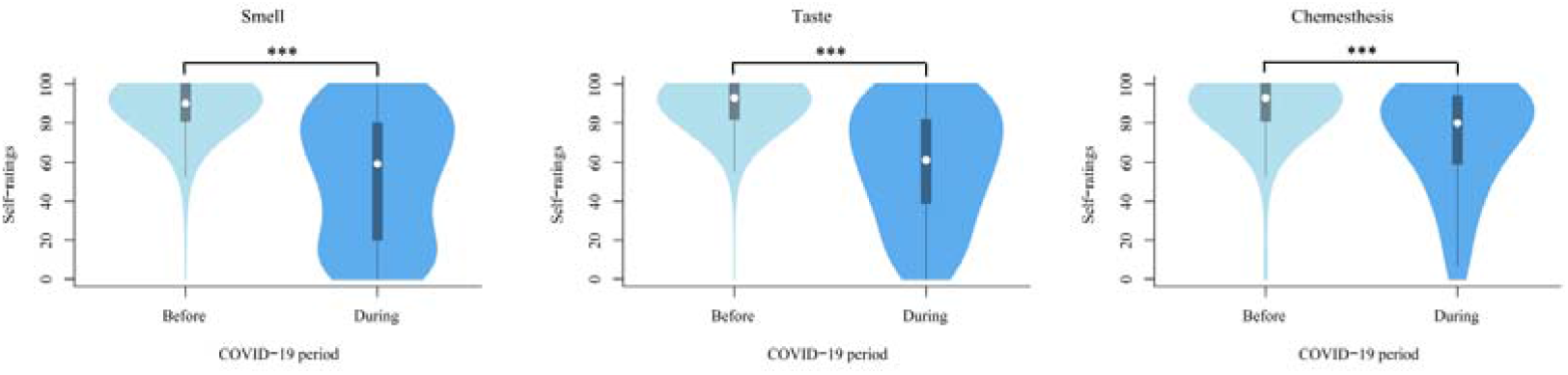
The results of self-rating of smell, taste, and chemesthesis function ^***^*p* < 0.001.

### Taste disorder

There were no statistically significant differences in demographic characteristics, such as sex, age group, and smoking/alcohol intake, between patients with and without taste disorder (all *p* > 0.05, Table 3). Patients with COVID-19-related symptoms, such as lack of appetite, dry and sore throat, myalgia, headache, diarrhea, cough/expectoration, stuffy/running nose, dyspnea, and fatigue had a significant chance of experiencing taste disorder (all p < 0.05). Associations between specific types of taste disorder (ageusia, hypogeusia, parageusia, phantogeusia) and demographic/COVID-19-related characteristics are detailed in the supplemental material (Table S6–S9). Additionally, of patients with total loss or decrease of specific taste (n = 597), 36.9% (220/597) reported total loss or decrease in the taste of sour, 45.9% (274/597) of sweet, 20.1% (120/597) of bitter, 51.4% (307/597) of salty, and 71.9% (429/597) of umami.

**Table 3.**
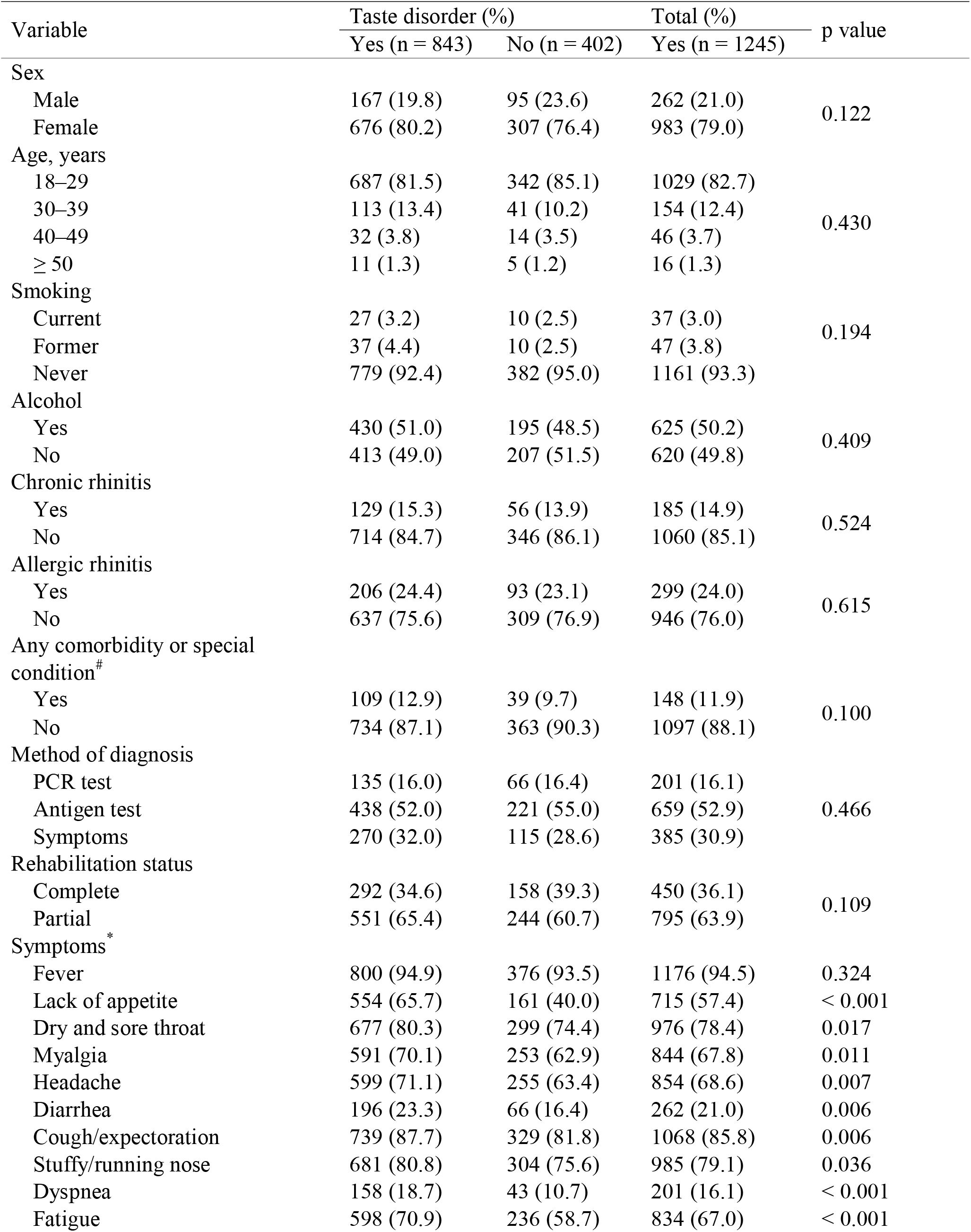
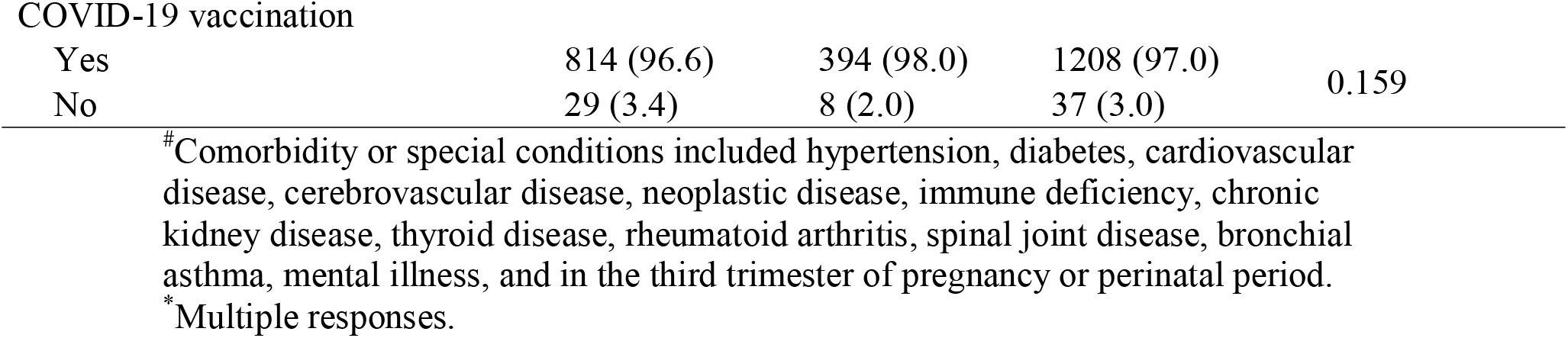
Association between taste disorder and demographic/COVID-19-related characteristics among patients.

| Variable | Taste disorder (%) |  | Total (%) | p value |
| --- | --- | --- | --- | --- |
|  | Yes (n = 843) | No (n = 402) | Yes (n = 1245) |  |
| Sex |  |  |  |  |
| Male | 167 (19.8) | 95 (23.6) | 262 (21.0) | 0.122 |
| Female | 676 (80.2) | 307 (76.4) | 983 (79.0) |  |
| Age, years |  |  |  |  |
| 18–29 | 687 (81.5) | 342 (85.1) | 1029 (82.7) | 0.430 |
| 30–39 | 113 (13.4) | 41 (10.2) | 154 (12.4) |  |
| 40–49 | 32 (3.8) | 14 (3.5) | 46 (3.7) |  |
| ≥ 50 | 11 (1.3) | 5 (1.2) | 16 (1.3) |  |
| Smoking |  |  |  |  |
| Current | 27 (3.2) | 10 (2.5) | 37 (3.0) | 0.194 |
| Former | 37 (4.4) | 10 (2.5) | 47 (3.8) |  |
| Never | 779 (92.4) | 382 (95.0) | 1161 (93.3) |  |
| Alcohol |  |  |  |  |
| Yes | 430 (51.0) | 195 (48.5) | 625 (50.2) | 0.409 |
| No | 413 (49.0) | 207 (51.5) | 620 (49.8) |  |
| Chronic rhinitis |  |  |  |  |
| Yes | 129 (15.3) | 56 (13.9) | 185 (14.9) | 0.524 |
| No | 714 (84.7) | 346 (86.1) | 1060 (85.1) |  |
| Allergic rhinitis |  |  |  |  |
| Yes | 206 (24.4) | 93 (23.1) | 299 (24.0) | 0.615 |
| No | 637 (75.6) | 309 (76.9) | 946 (76.0) |  |
| Any comorbidity or special condition<sup>#</sup> |  |  |  |  |
| Yes | 109 (12.9) | 39 (9.7) | 148 (11.9) | 0.100 |
| No | 734 (87.1) | 363 (90.3) | 1097 (88.1) |  |
| Method of diagnosis |  |  |  |  |
| PCR test | 135 (16.0) | 66 (16.4) | 201 (16.1) | 0.466 |
| Antigen test | 438 (52.0) | 221 (55.0) | 659 (52.9) |  |
| Symptoms | 270 (32.0) | 115 (28.6) | 385 (30.9) |  |
| Rehabilitation status |  |  |  |  |
| Complete | 292 (34.6) | 158 (39.3) | 450 (36.1) | 0.109 |
| Partial | 551 (65.4) | 244 (60.7) | 795 (63.9) |  |
| Symptoms<sup>*</sup> |  |  |  |  |
| Fever | 800 (94.9) | 376 (93.5) | 1176 (94.5) | 0.324 |
| Lack of appetite | 554 (65.7) | 161 (40.0) | 715 (57.4) | < 0.001 |
| Dry and sore throat | 677 (80.3) | 299 (74.4) | 976 (78.4) | 0.017 |
| Myalgia | 591 (70.1) | 253 (62.9) | 844 (67.8) | 0.011 |
| Headache | 599 (71.1) | 255 (63.4) | 854 (68.6) | 0.007 |
| Diarrhea | 196 (23.3) | 66 (16.4) | 262 (21.0) | 0.006 |
| Cough/expectorations | 739 (87.7) | 329 (81.8) | 1068 (85.8) | 0.006 |
| Stuffy/running nose | 681 (80.8) | 304 (75.6) | 985 (79.1) | 0.036 |
| Dyspnea | 158 (18.7) | 43 (10.7) | 201 (16.1) | < 0.001 |
| Fatigue | 598 (70.9) | 236 (58.7) | 834 (67.0) | < 0.001 |
| COVID-19 vaccination |  |  |  |  |
| Yes | 814 (96.6) | 394 (98.0) | 1208 (97.0) | 0.159 |
| No | 29 (3.4) | 8 (2.0) | 37 (3.0) |  |

Parageusia was described in 22.7% (282/1245) of patients, while phantogeusia was identified in 26.7% (333/1245) of patients. Pleasantness rating score on 100-point scale of the distorted gustatory sense caused by parosmia was 19.80 ± 21.33 (Mean ± SD), while that for gustatory hallucination was 24.97 ± 34.38 (Mean ± SD). Most patients with parageusia described that the flavors of foods, such as orange, meat, or candy, became bitter (e.g., “The rice has turned bitter”). Patients with phantogeusia most frequently reported a constant bitter flavor, without anything in mouth. Similar to smell, patients rated their taste function before and during COVID-19. The score during COVID-19 (58.78 ± 29.74, Mean ± SD) was significantly lower than that before COVID-19 (90.13 ± 11.60, Mean ± SD) (t = 37.173, p < 0.001).

### Chemesthesis disorder

No significant differences in demographic characteristics, including sex, age group, smoking and alcohol intake, chronic rhinitis, allergic rhinitis, and any comorbidity or special condition, were found between patients with and without chemesthesis disorder (all *p* > 0.05, Table 4). Significant differences in COVID-19-related symptoms (lack of appetite, dyspnea, and fatigue) were detected between patients with and without chemesthesis disorder (all *p* < 0.05).

**Table 4.**
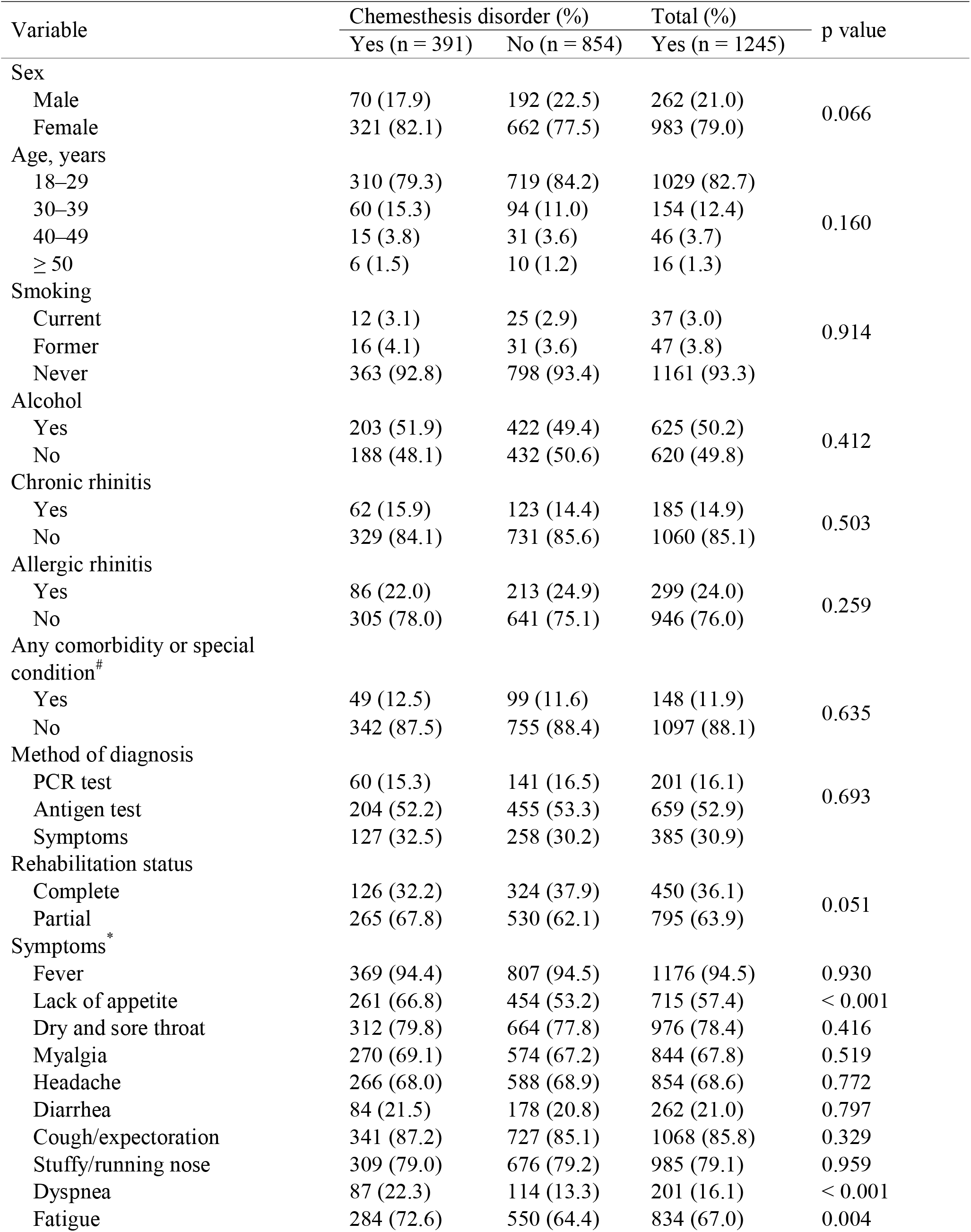
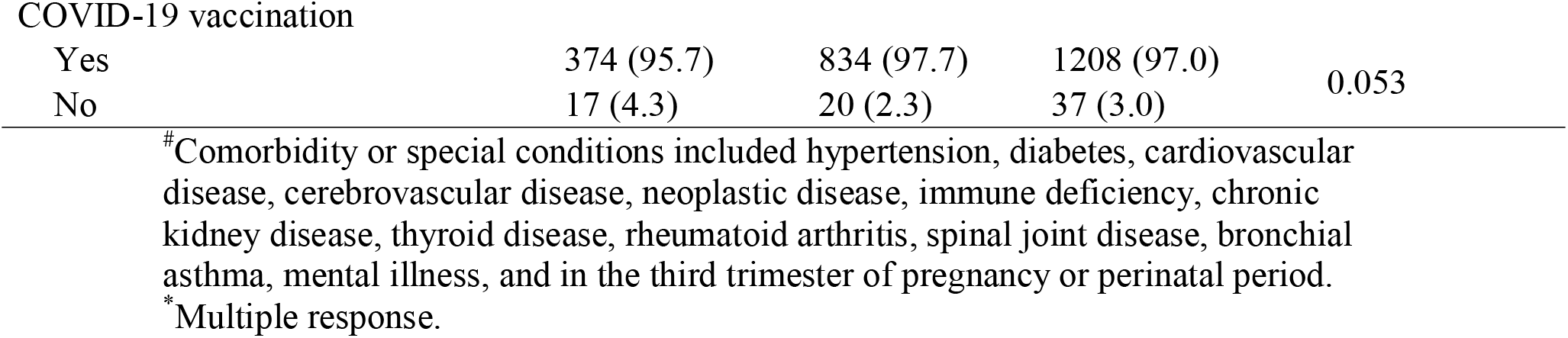
Association between chemesthesis disorder and patient demographic characteristics.

| Variable | Chemesthesis disorder (%) |  | Total (%) | p value |
| --- | --- | --- | --- | --- |
|  | Yes (n = 391) | No (n = 854) | Yes (n = 1245) |  |
| Sex |  |  |  |  |
| Male | 70 (17.9) | 192 (22.5) | 262 (21.0) | 0.066 |
| Female | 321 (82.1) | 662 (77.5) | 983 (79.0) |  |
| Age, years |  |  |  |  |
| 18–29 | 310 (79.3) | 719 (84.2) | 1029 (82.7) | 0.160 |
| 30–39 | 60 (15.3) | 94 (11.0) | 154 (12.4) |  |
| 40–49 | 15 (3.8) | 31 (3.6) | 46 (3.7) |  |
| ≥ 50 | 6 (1.5) | 10 (1.2) | 16 (1.3) |  |
| Smoking |  |  |  |  |
| Current | 12 (3.1) | 25 (2.9) | 37 (3.0) | 0.914 |
| Former | 16 (4.1) | 31 (3.6) | 47 (3.8) |  |
| Never | 363 (92.8) | 798 (93.4) | 1161 (93.3) |  |
| Alcohol |  |  |  |  |
| Yes | 203 (51.9) | 422 (49.4) | 625 (50.2) | 0.412 |
| No | 188 (48.1) | 432 (50.6) | 620 (49.8) |  |
| Chronic rhinitis |  |  |  |  |
| Yes | 62 (15.9) | 123 (14.4) | 185 (14.9) | 0.503 |
| No | 329 (84.1) | 731 (85.6) | 1060 (85.1) |  |
| Allergic rhinitis |  |  |  |  |
| Yes | 86 (22.0) | 213 (24.9) | 299 (24.0) | 0.259 |
| No | 305 (78.0) | 641 (75.1) | 946 (76.0) |  |
| Any comorbidity or special condition<sup>#</sup> |  |  |  |  |
| Yes | 49 (12.5) | 99 (11.6) | 148 (11.9) | 0.635 |
| No | 342 (87.5) | 755 (88.4) | 1097 (88.1) |  |
| Method of diagnosis |  |  |  |  |
| PCR test | 60 (15.3) | 141 (16.5) | 201 (16.1) | 0.693 |
| Antigen test | 204 (52.2) | 455 (53.3) | 659 (52.9) |  |
| Symptoms | 127 (32.5) | 258 (30.2) | 385 (30.9) |  |
| Rehabilitation status |  |  |  |  |
| Complete | 126 (32.2) | 324 (37.9) | 450 (36.1) | 0.051 |
| Partial | 265 (67.8) | 530 (62.1) | 795 (63.9) |  |
| Symptoms<sup>*</sup> |  |  |  |  |
| Fever | 369 (94.4) | 807 (94.5) | 1176 (94.5) | 0.930 |
| Lack of appetite | 261 (66.8) | 454 (53.2) | 715 (57.4) | < 0.001 |
| Dry and sore throat | 312 (79.8) | 664 (77.8) | 976 (78.4) | 0.416 |
| Myalgia | 270 (69.1) | 574 (67.2) | 844 (67.8) | 0.519 |
| Headache | 266 (68.0) | 588 (68.9) | 854 (68.6) | 0.772 |
| Diarrhea | 84 (21.5) | 178 (20.8) | 262 (21.0) | 0.797 |
| Cough/expectorations | 341 (87.2) | 727 (85.1) | 1068 (85.8) | 0.329 |
| Stuffy/running nose | 309 (79.0) | 676 (79.2) | 985 (79.1) | 0.959 |
| Dyspnea | 87 (22.3) | 114 (13.3) | 201 (16.1) | < 0.001 |
| Fatigue | 284 (72.6) | 550 (64.4) | 834 (67.0) | 0.004 |
| COVID-19 vaccination |  |  |  |  |
| Yes | 374 (95.7) | 834 (97.7) | 1208 (97.0) | 0.053 |
| No | 17 (4.3) | 20 (2.3) | 37 (3.0) |  |

Patients also rated their chemesthesis function before and during COVID-19. The score during COVID-19 (71.82 ± 26.48, Mean ± SD) was also significantly lower than that before COVID-19 (88.50 ± 15.83, Mean ± SD) (t = 24.700, p < 0.001).

## DISCUSSION

In the current study, we found that the prevalence rates of smell, taste, and chemesthesis disorder in 1245 patients with COVID-19 during the period of Omicron variant dominance were 69.2%, 67.7%, and 31.4%, respectively. Previous studies reported prevalence rates of olfactory dysfunction ranging from 3.2% to 98.3% and of taste dysfunction ranging from 5.6% to 62.7% ^(28,29)^. Previous research has found that the reported prevalence of smell, taste, and chemesthesis disorder varies according to differences in population, assessment method, and virus strain ^(28)^. A meta-analysis based on data from 24 studies, including over 8400 participants from 13 countries, found the pooled prevalence rates of patients with smell and taste dysfunction were 41.0% and 38.2%, respectively ^(29)^. Another meta-analysis involving 18 studies showed that the prevalence of alteration of the sense of smell or taste was 47% ^(30)^. Importantly, recent studies have focused primarily on quantitative changes in smell and taste, while qualitative changes of smell and taste were not addressed ^(29,31–33)^; however, we included five types of smell disorder (anosmia, hyposmia, parosmia, phantosmia and olfactory fluctuation), and four types of taste disorder (ageusia, hypogeusia, parageusia, phantogeusia) in our study. The prevalence of anosmia in our study was 23.3%, that of hyposmia was 42.5%, ageusia was 19.0%, and hypogeusia was 40.1%. Studies have also found that the prevalence of smell and taste disorder decreased during the period of Omicron variant dominance, but remained more than 30%, similar to our findings ^(28,34)^.

Males are reported to be more prone to loss of smell and taste senses than females ^(35)^; however, our data demonstrate that females were more prone to complete loss of olfaction, similar to the findings of Lechien et al. ^(6,8)^. In terms of taste loss, we did not detect significant differences between the sexes, similar to Al-Rawi et al. ^(3,36)^. Further, Al-Rawi et al. found more significant smell and taste loss among individuals in their late 30s ^(3)^, consistent with our finding that patients aged 30–39 years are more prone to anosmia and ageusia. Previous research showed that smokers are more vulnerable to anosmia and ageusia ^(35)^; however, we detected no significant differences between smokers and non-smokers in terms of anosmia and ageusia in our study. Interestingly, we found that smoking had adverse effects on smell disorders in general.

We also found that some COVID-19 related symptoms, such as lack of appetite, dyspnea, and fatigue, were associated with chemosensory disorder rates. Associations of these factors with chemosensory disorders have been reported in previous research ^(7,8)^; however, whether they are risk factors for chemosensory disorders requires further study.

In the current study, we report the prevalence, quality, and quantity of chemosensory changes in patients infected with the Omicron variant in China. Patients often confuse chemesthesis with smell and taste when self-reporting, thus our survey included an expanded section relating to chemesthesis, with the aim of better distinguishing the influences of COVID-19 infection on smell, taste and chemesthesis. We also made detailed distinctions among the quality and quantity of changes in smell and taste, and found that these changes differed in the COVID-19 infected population, which may provide reference data for the follow-up research on the mechanism underlying COVID-19 influence on smell and taste. Moreover, we expanded the Chinese version of the GCCR core questionnaire, to provide reference data for future domestic research.

Our study also had several limitations. First, the participants were mostly from the southern areas of China. Second, the collection of the data for our study was based on self-report, which may lead to confirmation bias of chemosensory disorders. Additional objective assessment of chemosensory disorders is needed in future studies. Third, our study was cross-sectional, and longitudinal studies are needed to determine the direction of associations among variables in our study.

In conclusion, the current study found that the prevalence rates of smell, taste, and chemesthesis disorder in 1245 patients with COVID-19 during the period of Omicron variant dominance were 69.2%, 67.7%, and 31.4%, respectively. Patients experienced a general decline in the function of smell, taste, and chemesthesis. Sex, age, smoking, and COVID-19-related symptoms, such as lack of appetite, dyspnea, and fatigue, may be associated with chemosensory disorders during COVID-19. The present study contributes to existing knowledge of chemosensory disorders in COVID-19 by providing Chinese data collected during Omicron variant dominance.

## Data Availability

All data produced in the present study are available upon reasonable request to the authors.

## ACKNOWLEDGEMENTS

We thank all the participants who participated in our study.

## AUTHORSHIP CONTRIBUTION

Conceptualization, Laiquan Zou; formal analysis, Ying Chen, Yuying Chen, and Laiquan Zou; investigation, Ying Chen, Yuying Chen, Xiang Liu, Chao Yan, and Laiquan Zou; writing−original draft, Ying Chen, Yuying Chen, and Laiquan Zou; writing—review & editing, Xiang Liu, Chao Yan, and Laiquan Zou. All authors have read and agreed to the published version of the manuscript.

