## Supplementary_Data for "Smell, taste and chemesthesis disorders in patients with the SARS-CoV-2 Omicron variant in China"

**S1 Association between anosmia and demographic/COVID-19-related characteristics among patients**

| Variables | | Anosmia (%) | | Total (%) | p value |
| --- | --- | --- | --- | --- | --- |
|  |  | Yes (n = 290) | No (n = 955) | N = 1245 |  |
| Sex | |  |  |  |  |
|  | Male | 49 (16.9) | 213 (22.3) | 262 (21.0) | 0.048 |
|  | Female | 241 (83.1) | 742 (77.7) | 983 (79.0) |  |
| Age, yr | |  |  |  |  |
|  | 18-29 | 232 (80.0) | 797 (83.5) | 1029 (82.7) | 0.044 |
|  | 30-39 | 47 (16.2) | 107 (11.2) | 154 (12.4) |  |
|  | 40-49 | 6 (2.1) | 40 (4.2) | 46 (3.7) |  |
|  | 50 | 5 (1.7) | 11 (1.2) | 16 (1.3) |  |
| Smoking | |  |  |  |  |
|  | Current | 14 (4.8) | 23 (2.4) | 37 (3.0) | 0.054 |
|  | Former | 14 (4.8) | 33 (3.5) | 47 (3.8) |  |
|  | Never | 262 (90.3) | 899 (94.1) | 1161 (93.3) |  |
| Alcohol | |  |  |  |  |
|  | Yes | 154 (53.1) | 471 (49.3) | 625 (50.2) | 0.259 |
|  | No | 136 (46.9) | 484 (50.7) | 620 (49.8) |  |
| Chronic rhinitis | |  |  |  |  |
|  | Yes | 41 (14.1) | 144 (15.1) | 185 (14.9) | 0.693 |
|  | No | 249 (85.9) | 811 (84.9) | 1060 (85.1) |  |
| Allergic rhinitis | |  |  |  |  |
|  | Yes | 70 (24.1) | 229 (24.0) | 299 (24.0) | 0.956 |
|  | No | 220 (75.9) | 726 (76.0) | 946 (76.0) |  |
| Any comorbidity or special condition# | |  |  |  |  |
|  | Yes | 39 (13.4) | 109 (11.4) | 148 (11.9) | 0.348 |
|  | No | 251 (86.6) | 846 (88.6) | 1097 (88.1) |  |
| Method of diagnosis | |  |  |  |  |
|  | PCR test | 46 (15.9) | 155 (16.2) | 201 (16.1) | 0.740 |
|  | Antigen test | 149 (51.4) | 510 (53.4) | 659 (52.9) |  |
|  | Symptoms | 95 (32.8) | 290 (30.4) | 385 (30.9) |  |
| Rehabilitation status | |  |  |  |  |
|  | Complete | 85 (29.3) | 365 (38.2) | 450 (36.1) | 0.006 |
|  | Partial | 205 (70.7) | 590 (61.8) | 795 (63.9) |  |
| Symptoms | |  |  |  |  |
|  | Fever | 278 (95.9) | 898 (94.0) | 1176 (94.5) | 0.233 |
|  | Lack of appetite | 184 (63.4) | 531 (55.6) | 715 (57.4) | 0.018 |
|  | Throat dryness and sore | 231 (79.7) | 745 (78.0) | 976 (78.4) | 0.551 |
|  | Myalgia | 212 (73.1) | 632 (66.2) | 844 (67.8) | 0.027 |
|  | Headache | 194 (66.9) | 660 (69.1) | 854 (68.6) | 0.477 |
|  | Diarrhea | 60 (20.7) | 202 (21.2) | 262 (21.0) | 0.866 |
|  | Cough/expectoration | 255 (87.9) | 813 (85.1) | 1068 (85.8) | 0.232 |
|  | Stuffy/running nose | 242 (83.4) | 743 (77.8) | 985 (79.1) | 0.038 |
|  | Dyspnea | 49 (16.9) | 152 (15.9) | 201 (16.1) | 0.691 |
|  | Fatigue | 202 (69.7) | 632 (66.2) | 834 (67.0) | 0.270 |
| COVID-19 vaccination | |  |  |  |  |
|  | Yes (n = 1208) | 273 (94.1) | 935 (97.9) | 1208 (97.0) | ＜0.001 |
|  | No (n = 37) | 17 (5.9) | 20 (2.1) | 37 (3.0) |  |

^#^ Comorbidity or special condition includes hypertension, diabetes, cardiovascular disease, cerebrovascular disease, neoplastic disease, immune deficiency, chronic kidney disease, thyroid disease, rheumatoid arthritis, spinal joint disease, bronchial asthma, mental illness, and the third pregnancy and perinatal period

^*^ Multiple response

**S2 Association between hyposmia and demographic/COVID-19-related characteristics among patients**

| Variables | | Hyposmia (%) | | Total (%) | p value |
| --- | --- | --- | --- | --- | --- |
|  |  | Yes (n = 529) | No (n = 716) | N = 1245 |  |
| Sex | |  |  |  |  |
|  | Male | 113 (21.4) | 149 (20.8) | 262 (21.0) | 0.814 |
|  | Female | 416 (78.6) | 567 (79.2) | 983 (79.0) |  |
| Age, yr | |  |  |  |  |
|  | 18-29 | 433 (81.9) | 596 (83.2) | 1029 (82.7) | 0.765 |
|  | 30-39 | 66 (12.5) | 88 (12.3) | 154 (12.4) |  |
|  | 40-49 | 23 (4.3) | 23 (3.2) | 46 (3.7) |  |
|  | ≥50 | 7 (1.3) | 9 (1.3) | 16 (1.3) |  |
| Smoking | |  |  |  |  |
|  | Current | 17 (3.2) | 20 (2.8) | 37 (3.0) | 0.593 |
|  | Former | 23 (4.3) | 24 (3.4) | 47 (3.8) |  |
|  | Never | 489 (92.4) | 672 (93.9) | 1161 (93.3) |  |
| Alcohol | |  |  |  |  |
|  | Yes | 270 (51.0) | 355 (49.6) | 625 (50.2) | 0.611 |
|  | No | 259 (49.0) | 361 (50.4) | 620 (49.8) |  |
| Chronic rhinitis | |  |  |  |  |
|  | Yes | 89 (16.8) | 96 (13.4) | 185 (14.9) | 0.094 |
|  | No | 440 (83.2) | 620 (86.6) | 1060 (85.1) |  |
| Allergic rhinitis | |  |  |  |  |
|  | Yes | 127 (24.0) | 172 (24.0) | 299 (24.0) | 0.995 |
|  | No | 402 (76.0) | 544 (76.0) | 946 (76.0) |  |
| Any comorbidity or special condition^#^ | |  |  |  |  |
|  | Yes | 61 (11.5) | 87 (12.2) | 148 (11.9) | 0.738 |
|  | No | 468 (88.5) | 629 (87.8) | 1097 (88.1) |  |
| Method of diagnosis | |  |  |  |  |
|  | PCR test | 83 (15.7) | 118 (16.5) | 201 (16.1) | 0.582 |
|  | Antigen test | 289 (54.6) | 370 (51.7) | 659 (52.9) |  |
|  | Symptoms | 157 (29.7) | 228 (31.8) | 385 (30.9) |  |
| Rehabilitation status | |  |  |  |  |
|  | Complete | 181 (34.2) | 269 (37.6) | 450 (36.1) | 0.223 |
|  | Partial | 348 (65.8) | 447 (62.4) | 795 (63.9) |  |
| Symptoms^*^ | |  |  |  |  |
|  | Fever | 501 (94.7) | 675 (94.3) | 1176 (94.5) | 0.741 |
|  | Lack of appetite | 341 (64.5) | 374 (52.2) | 715 (57.4) | < 0.001 |
|  | Throat dryness and sore | 435 (82.2) | 541 (75.6) | 976 (78.4) | 0.005 |
|  | Myalgia | 365 (69.0) | 479 (66.9) | 844 (67.8) | 0.433 |
|  | Headache | 381 (72.0) | 473 (66.1) | 854 (68.6) | 0.025 |
|  | Diarrhea | 126 (23.8) | 136 (19.0) | 262 (21.0) | 0.039 |
|  | Cough/expectoration | 459 (86.8) | 609 (85.1) | 1068 (85.8) | 0.393 |
|  | Stuffy/running nose | 431 (81.5) | 554 (77.4) | 985 (79.1) | 0.079 |
|  | Dyspnea | 100 (18.9) | 101 (14.1) | 201 (16.1) | 0.023 |
|  | Fatigue | 372 (70.3) | 462 (64.5) | 834 (67.0) | 0.032 |
| COVID-19 vaccination | |  |  |  |  |
|  | Yes (n = 1208) | 514 (97.2) | 694 (96.9) | 1208 (97.0) | 0.808 |
|  | No (n = 37) | 15 (2.8) | 22 (3.1) | 37 (3.0) |  |

^#^ Comorbidity or special condition includes hypertension, diabetes, cardiovascular disease, cerebrovascular disease, neoplastic disease, immune deficiency, chronic kidney disease, thyroid disease, rheumatoid arthritis, spinal joint disease, bronchial asthma, mental illness, and the third pregnancy and perinatal period

^*^ Multiple response

**S3 Association between** **paraosmia and demographic/COVID-19-related characteristics among patients**

| Variables | | Paraosmia (%) | | Total (%) | p value |
| --- | --- | --- | --- | --- | --- |
|  |  | Yes (n = 60) | No (n = 1185) | N = 1245 |  |
| Sex | |  |  |  |  |
|  | Male | 14 (23.3) | 248 (20.9) | 262 (21.0) | 0.656 |
|  | Female | 46 (76.7) | 937 (79.1) | 983 (79.0) |  |
| Age, yr | |  |  |  |  |
|  | 18-29 | 42 (70.0) | 987 (83.3) | 1029 (82.7) | 0.003 |
|  | 30-39 | 12 (20.0) | 142 (12.0) | 154 (12.4) |  |
|  | 40-49 | 2 (3.3) | 44 (3.7) | 46 (3.7) |  |
|  | ≥50 | 4 (6.7) | 12 (1.0) | 16 (1.3) |  |
| Smoking | |  |  |  |  |
|  | Current | 5 (8.3) | 32 (2.7) | 37 (3.0) | 0.007 |
|  | Former | 5 (8.3) | 42 (3.5) | 47 (3.8) |  |
|  | Never | 50 (83.3) | 1111 (93.8) | 1161 (93.3) |  |
| Alcohol | |  |  |  |  |
|  | Yes | 32 (53.3) | 593 (50.0) | 625 (50.2) | 0.619 |
|  | No | 28 (46.7) | 592 (50.0) | 620 (49.8) |  |
| Chronic rhinitis | |  |  |  |  |
|  | Yes | 12 (20.0) | 173 (14.6) | 185 (14.9) | 0.251 |
|  | No | 48 (80.0) | 1012 (85.4) | 1060 (85.1) |  |
| Allergic rhinitis | |  |  |  |  |
|  | Yes | 18 (30.0) | 281 (23.7) | 299 (24.0) | 0.266 |
|  | No | 42 (70.0) | 904 (76.3) | 946 (76.0) |  |
| Any comorbidity or special condition^#^ | |  |  |  |  |
|  | Yes | 7 (11.7) | 141 (11.9) | 148 (11.9) | 0.957 |
|  | No | 53 (88.3) | 1044 (88.1) | 1097 (88.1) |  |
| Method of diagnosis | |  |  |  |  |
|  | PCR test | 10 (16.7) | 191 (16.1) | 201 (16.1) | 0.891 |
|  | Antigen test | 30 (50.0) | 629 (53.1) | 659 (52.9) |  |
|  | Symptoms | 20 (33.3) | 365 (30.8) | 385 (30.9) |  |
| Rehabilitation status | |  |  |  |  |
|  | Complete | 17 (28.3) | 433 (36.5) | 450 (36.1) | 0.197 |
|  | Partial | 43 (71.7) | 752 (63.5) | 795 (63.9) |  |
| Symptoms^*^ | |  |  |  |  |
|  | Fever | 57 (95.0) | 1119 (94.4) | 1176 (94.5) | 1.000 |
|  | Lack of appetite | 39 (65.0) | 676 (57.0) | 715 (57.4) | 0.224 |
|  | Throat dryness and sore | 51 (85.0) | 925 (78.1) | 976 (78.4) | 0.202 |
|  | Myalgia | 42 (70.0) | 802 (67.7) | 844 (67.8) | 0.707 |
|  | Headache | 49 (81.7) | 805 (67.9) | 854 (68.6) | 0.025 |
|  | Diarrhea | 19 (31.7) | 243 (20.5) | 262 (21.0) | 0.039 |
|  | Cough/expectoration | 50 (83.3) | 1018 (85.9) | 1068 (85.8) | 0.578 |
|  | Stuffy/running nose | 49 (81.7) | 936 (79.0) | 985 (79.1) | 0.618 |
|  | Dyspnea | 13 (21.7) | 188 (15.9) | 201 (16.1) | 0.233 |
|  | Fatigue | 46 (76.7) | 788 (66.5) | 834 (67.0) | 0.102 |
| COVID-19 vaccination | |  |  |  |  |
|  | Yes (n = 1208) | 59 (98.3) | 1149 (97.0) | 1208 (97.0) | 1.000 |
|  | No (n = 37) | 1 (1.7) | 36 (3.0) | 37 (3.0) |  |

^#^ Comorbidity or special condition includes hypertension, diabetes, cardiovascular disease, cerebrovascular disease, neoplastic disease, immune deficiency, chronic kidney disease, thyroid disease, rheumatoid arthritis, spinal joint disease, bronchial asthma, mental illness, and the third pregnancy and perinatal period

^*^ Multiple response

**S4 Association between** **phantosmia and demographic/COVID-19-related characteristics among patients**

| Variables | | Phantosmia (%) | | Total (%) | p value |
| --- | --- | --- | --- | --- | --- |
|  |  | Yes (n = 98) | No (n = 1147) | N = 1245 |  |
| Sex | |  |  |  |  |
|  | Male | 17 (17.3) | 245 (21.4) | 262 (21.0) | 0.350 |
|  | Female | 81 (82.7) | 902 (78.6) | 983 (79.0) |  |
| Age, yr | |  |  |  |  |
|  | 18-29 | 83 (84.7) | 946 (82.5) | 1029 (82.7) | 0.669 |
|  | 30-39 | 11 (11.2) | 143 (12.5) | 154 (12.4) |  |
|  | 40-49 | 2 (2.0) | 44 (3.8) | 46 (3.7) |  |
|  | ≥50 | 2 (2.0) | 14 (1.2) | 16 (1.3) |  |
| Smoking | |  |  |  |  |
|  | Current | 2 (2.0) | 35 (3.1) | 37 (3.0) | 0.413 |
|  | Former | 6 (6.1) | 41 (3.6) | 47 (3.8) |  |
|  | Never | 90 (91.8) | 1071 (93.4) | 1161 (93.3) |  |
| Alcohol | |  |  |  |  |
|  | Yes | 51 (52.0) | 574 (50.0) | 625 (50.2) | 0.704 |
|  | No | 47 (48.0) | 573 (50.0) | 620 (49.8) |  |
| Chronic rhinitis | |  |  |  |  |
|  | Yes | 18 (18.4) | 167 (14.6) | 185 (14.9) | 0.309 |
|  | No | 80 (81.6) | 980 (85.4) | 1060 (85.1) |  |
| Allergic rhinitis | |  |  |  |  |
|  | Yes | 30 (30.6) | 269 (23.5) | 299 (24.0) | 0.111 |
|  | No | 68 (69.4) | 878 (76.5) | 946 (76.0) |  |
| Any comorbidity or special condition^#^ | |  |  |  |  |
|  | Yes | 14 (14.3) | 134 (11.7) | 148 (11.9) | 0.445 |
|  | No | 84 (85.7) | 1013 (88.3) | 1097 (88.1) |  |
| Method of diagnosis | |  |  |  |  |
|  | PCR test | 13 (13.3) | 188 (16.4) | 201 (16.1) | 0.591 |
|  | Antigen test | 51 (52.0) | 608 (53.0) | 659 (52.9) |  |
|  | Symptoms | 34 (34.7) | 351 (30.6) | 385 (30.9) |  |
| Rehabilitation status | |  |  |  |  |
|  | Complete | 29 (29.6) | 421 (36.7) | 450 (36.1) | 0.159 |
|  | Partial | 69 (70.4) | 726 (63.3) | 795 (63.9) |  |
| Symptoms^*^ | |  |  |  |  |
|  | Fever | 94 (95.9) | 1082 (94.3) | 1176 (94.5) | 0.510 |
|  | Lack of appetite | 60 (61.2) | 655 (57.1) | 715 (57.4) | 0.429 |
|  | Throat dryness and sore | 72 (73.5) | 904 (78.8) | 976 (78.4) | 0.217 |
|  | Myalgia | 69 (70.4) | 775 (67.6) | 844 (67.8) | 0.564 |
|  | Headache | 65 (66.3) | 789 (68.8) | 854 (68.6) | 0.614 |
|  | Diarrhea | 28 (28.6) | 234 (20.4) | 262 (21.0) | 0.057 |
|  | Cough/expectoration | 77 (78.6) | 991 (86.4) | 1068 (85.8) | 0.033 |
|  | Stuffy/running nose | 78 (79.6) | 907 (79.1) | 985 (79.1) | 0.904 |
|  | Dyspnea | 24 (24.5) | 177 (15.4) | 201 (16.1) | 0.019 |
|  | Fatigue | 72 (73.5) | 762 (66.4) | 834 (67.0) | 0.155 |
| COVID-19 vaccination | |  |  |  |  |
|  | Yes (n = 1208) | 97 (99.0) | 1111 (96.9) | 1208 (97.0) | 0.356 |
|  | No (n = 37) | 1 (1.0) | 36 (3.1) | 37 (3.0) |  |

^#^ Comorbidity or special condition includes hypertension, diabetes, cardiovascular disease, cerebrovascular disease, neoplastic disease, immune deficiency, chronic kidney disease, thyroid disease, rheumatoid arthritis, spinal joint disease, bronchial asthma, mental illness, and the third pregnancy and perinatal period

^*^ Multiple response

**S5 Association between** **olfactory fluctuation and demographic/COVID-19-related characteristics among patients**

| Variables | | Olfactory fluctuation (%) | | Total (%) | p value |
| --- | --- | --- | --- | --- | --- |
|  |  | Yes (n = 438) | No (n = 807) | N = 1245 |  |
| Sex | |  |  |  |  |
|  | Male | 66 (15.1) | 196 (24.3) | 262 (21.0) | <0.001 |
|  | Female | 372 (84.9) | 611 (75.7) | 983 (79.0) |  |
| Age, yr | |  |  |  |  |
|  | 18-29 | 379 (86.5) | 650 (80.5) | 1029 (82.7) | 0.059 |
|  | 30-39 | 43 (9.8) | 111 (13.8) | 154 (12.4) |  |
|  | 40-49 | 11 (2.5) | 35 (4.3) | 46 (3.7) |  |
|  | ≥50 | 5 (1.1) | 11 (1.4) | 16 (1.3) |  |
| Smoking | |  |  |  |  |
|  | Current | 15 (3.4) | 22 (2.7) | 37 (3.0) | 0.779 |
|  | Former | 16 (3.7) | 31 (3.8) | 47 (3.8) |  |
|  | Never | 407 (92.9) | 754 (93.4) | 1161 (93.3) |  |
| Alcohol | |  |  |  |  |
|  | Yes | 230 (52.5) | 395 (48.9) | 625 (50.2) | 0.230 |
|  | No | 208 (47.5) | 412 (51.1) | 620 (49.8) |  |
| Chronic rhinitis | |  |  |  |  |
|  | Yes | 75 (17.1) | 110 (13.6) | 185 (14.9) | 0.098 |
|  | No | 363 (82.9) | 697 (86.4) | 1060 (85.1) |  |
| Allergic rhinitis | |  |  |  |  |
|  | Yes | 106 (24.2) | 193 (23.9) | 299 (24.0) | 0.910 |
|  | No | 332 (75.8) | 614 (76.1) | 946 (76.0) |  |
| Any comorbidity or special condition^#^ | |  |  |  |  |
|  | Yes | 50 (11.4) | 98 (12.1) | 148 (11.9) | 0.705 |
|  | No | 388 (88.6) | 709 (87.9) | 1097 (88.1) |  |
| Method of diagnosis | |  |  |  |  |
|  | PCR test | 71 (16.2) | 130 (16.1) | 201 (16.1) | 0.885 |
|  | Antigen test | 228 (52.1) | 431 (53.4) | 659 (52.9) |  |
|  | Symptoms | 139 (31.7) | 246 (30.5) | 385 (30.9) |  |
| Rehabilitation status | |  |  |  |  |
|  | Complete | 142 (32.4) | 308 (38.2) | 450 (36.1) | 0.044 |
|  | Partial | 296 (67.6) | 499 (61.8) | 795 (63.9) |  |
| Symptoms^*^ | |  |  |  |  |
|  | Fever | 420 (95.9) | 756 (93.7) | 1176 (94.5) | 0.104 |
|  | Lack of appetite | 267 (61.0) | 448 (55.5) | 715 (57.4) | 0.064 |
|  | Throat dryness and sore | 361 (82.4) | 615 (76.2) | 976 (78.4) | 0.011 |
|  | Myalgia | 317 (72.4) | 527 (65.3) | 844 (67.8) | 0.011 |
|  | Headache | 313 (71.5) | 541 (67.0) | 854 (68.6) | 0.108 |
|  | Diarrhea | 92 (21.0) | 170 (21.1) | 262 (21.0) | 0.980 |
|  | Cough/expectoration | 384 (87.7) | 684 (84.8) | 1068 (85.8) | 0.160 |
|  | Stuffy/running nose | 356 (81.3) | 629 (77.9) | 985 (79.1) | 0.167 |
|  | Dyspnea | 90 (20.5) | 111 (13.8) | 201 (16.1) | 0.002 |
|  | Fatigue | 315 (71.9) | 519 (64.3) | 834 (67.0) | 0.006 |
| COVID-19 vaccination | |  |  |  |  |
|  | Yes (n = 1208) | 424 (96.8) | 784 (97.1) | 1208 (97.0) | 0.731 |
|  | No (n = 37) | 14 (3.2) | 23 (2.9) | 37 (3.0) |  |

^#^ Comorbidity or special condition includes hypertension, diabetes, cardiovascular disease, cerebrovascular disease, neoplastic disease, immune deficiency, chronic kidney disease, thyroid disease, rheumatoid arthritis, spinal joint disease, bronchial asthma, mental illness, and the third pregnancy and perinatal period

^*^ Multiple response

**S6 Association between** **ageusia and demographic/COVID-19-related characteristics among patients**

| Variables | | Ageusia (%) | | Total (%) | p value |
| --- | --- | --- | --- | --- | --- |
|  |  | Yes (n = 236) | No (n = 1009) | N = 1245 |  |
| Sex | |  |  |  |  |
|  | Male | 44 (18.6) | 218 (21.6) | 262 (21.0) | 0.315 |
|  | Female | 192 (81.4) | 791 (78.4) | 983 (79.0) |  |
| Age, yr | |  |  |  |  |
|  | 18-29 | 178 (75.4) | 851 (84.3) | 1029 (82.7) | 0.005 |
|  | 30-39 | 44 (18.6) | 110 (10.9) | 154 (12.4) |  |
|  | 40-49 | 9 (3.8) | 37 (3.7) | 46 (3.7) |  |
|  | ≥50 | 5 (2.1) | 11 (1.1) | 16 (1.3) |  |
| Smoking | |  |  |  |  |
|  | Current | 7 (3.0) | 30 (3.0) | 37 (3.0) | 0.918 |
|  | Former | 10 (4.2) | 37 (3.7) | 47 (3.8) |  |
|  | Never | 219 (92.8) | 942 (93.4) | 1161 (93.3) |  |
| Alcohol | |  |  |  |  |
|  | Yes | 118 (50.0) | 507 (50.2) | 625 (50.2) | 0.945 |
|  | No | 118 (50.0) | 502 (49.8) | 620 (49.8) |  |
| Chronic rhinitis | |  |  |  |  |
|  | Yes | 35 (14.8) | 150 (14.9) | 185 (14.9) | 0.989 |
|  | No | 201 (85.2) | 859 (85.1) | 1060 (85.1) |  |
| Allergic rhinitis | |  |  |  |  |
|  | Yes | 58 (24.6) | 241 (23.9) | 299 (24.0) | 0.823 |
|  | No | 178 (75.4) | 768 (76.1) | 946 (76.0) |  |
| Any comorbidity or special condition^#^ | |  |  |  |  |
|  | Yes | 38 (16.1) | 110 (10.9) | 148 (11.9) | 0.026 |
|  | No | 198 (83.9) | 899 (89.1) | 1097 (88.1) |  |
| Method of diagnosis | |  |  |  |  |
|  | PCR test | 33 (14.0) | 168 (16.7) | 201 (16.1) | 0.530 |
|  | Antigen test | 125 (53.0) | 534 (52.9) | 659 (52.9) |  |
|  | Symptoms | 78 (33.1) | 307 (30.4) | 385 (30.9) |  |
| Rehabilitation status | |  |  |  |  |
|  | Complete | 71 (30.1) | 379 (37.6) | 450 (36.1) | 0.031 |
|  | Partial | 165 (69.9) | 630 (62.4) | 795 (63.9) |  |
| Symptoms^*^ | |  |  |  |  |
|  | Fever | 221 (93.6) | 955 (94.6) | 1176 (94.5) | 0.544 |
|  | Lack of appetite | 144 (61.0) | 571 (56.6) | 715 (57.4) | 0.216 |
|  | Throat dryness and sore | 190 (80.5) | 786 (77.9) | 976 (78.4) | 0.381 |
|  | Myalgia | 167 (70.8) | 677 (67.1) | 844 (67.8) | 0.278 |
|  | Headache | 162 (68.6) | 692 (68.6) | 854 (68.6) | 0.985 |
|  | Diarrhea | 58 (24.6) | 204 (20.2) | 262 (21.0) | 0.139 |
|  | Cough/expectoration | 204 (86.4) | 864 (85.6) | 1068 (85.8) | 0.748 |
|  | Stuffy/running nose | 191 (80.9) | 794 (78.7) | 985 (79.1) | 0.446 |
|  | Dyspnea | 50 (21.2) | 151 (15.0) | 201 (16.1) | 0.019 |
|  | Fatigue | 167 (70.8) | 667 (66.1) | 834 (67.0) | 0.171 |
| COVID-19 vaccination | |  |  |  |  |
|  | Yes (n = 1208) | 228 (96.6) | 980 (97.1) | 1208 (97.0) | 0.674 |
|  | No (n = 37) | 8 (3.4) | 29 (2.9) | 37 (3.0) |  |

^#^ Comorbidity or special condition includes hypertension, diabetes, cardiovascular disease, cerebrovascular disease, neoplastic disease, immune deficiency, chronic kidney disease, thyroid disease, rheumatoid arthritis, spinal joint disease, bronchial asthma, mental illness, and the third pregnancy and perinatal period

^*^ Multiple response

**S7 Association between** **hypogeusia and demographic/COVID-19-related characteristics among patients**

| Variables | | Hypogeusia (%) | | Total (%) | p value |
| --- | --- | --- | --- | --- | --- |
|  |  | Yes (n = 499) | No (n = 746) | N = 1245 |  |
| Sex | |  |  |  |  |
|  | Male | 109 (21.8) | 153 (20.5) | 262 (21.0) | 0.571 |
|  | Female | 390 (78.2) | 593 (79.5) | 983 (79.0) |  |
| Age, yr | |  |  |  |  |
|  | 18-29 | 417 (83.6) | 612 (82.0) | 1029 (82.7) | 0.252 |
|  | 30-39 | 58 (11.6) | 96 (12.9) | 154 (12.4) |  |
|  | 40-49 | 21 (4.2) | 25 (3.4) | 46 (3.7) |  |
|  | ≥50 | 3 (0.6) | 13 (1.7) | 16 (1.3) |  |
| Smoking | |  |  |  |  |
|  | Current | 18 (3.6) | 19 (2.5) | 37 (3.0) | 0.439 |
|  | Former | 21 (4.2) | 26 (3.5) | 47 (3.8) |  |
|  | Never | 460 (92.2) | 701 (94.0) | 1161 (93.3) |  |
| Alcohol | |  |  |  |  |
|  | Yes | 258 (51.7) | 367 (49.2) | 625 (50.2) | 0.386 |
|  | No | 241 (48.3) | 379 (50.8) | 620 (49.8) |  |
| Chronic rhinitis | |  |  |  |  |
|  | Yes | 80 (16.0) | 105 (14.1) | 185 (14.9) | 0.341 |
|  | No | 419 (84.0) | 641 (85.9) | 1060 (85.1) |  |
| Allergic rhinitis | |  |  |  |  |
|  | Yes | 122 (24.4) | 177 (23.7) | 299 (24.0) | 0.770 |
|  | No | 377 (75.6) | 569 (76.3) | 946 (76.0) |  |
| Any comorbidity or special condition^#^ | |  |  |  |  |
|  | Yes | 57 (11.4) | 91 (12.2) | 148 (11.9) | 0.679 |
|  | No | 442 (88.6) | 655 (87.8) | 1097 (88.1) |  |
| Method of diagnosis | |  |  |  |  |
|  | PCR test | 88 (17.6) | 113 (15.1) | 201 (16.1) | 0.456 |
|  | Antigen test | 256 (51.3) | 403 (54.0) | 659 (52.9) |  |
|  | Symptoms | 155 (31.1) | 230 (30.8) | 385 (30.9) |  |
| Rehabilitation status | |  |  |  |  |
|  | Complete | 178 (35.7) | 272 (36.5) | 450 (36.1) | 0.776 |
|  | Partial | 321 (64.3) | 474 (63.5) | 795 (63.9) |  |
| Symptoms^*^ | |  |  |  |  |
|  | Fever | 475 (95.2) | 701 (94.0) | 1176 (94.5) | 0.356 |
|  | Lack of appetite | 345 (69.1) | 370 (49.6) | 715 (57.4) | < 0.001 |
|  | Throat dryness and sore | 400 (80.2) | 576 (77.2) | 976 (78.4) | 0.215 |
|  | Myalgia | 342 (68.5) | 502 (67.3) | 844 (67.8) | 0.645 |
|  | Headache | 357 (71.5) | 497 (66.6) | 854 (68.6) | 0.067 |
|  | Diarrhea | 113 (22.6) | 149 (20.0) | 262 (21.0) | 0.257 |
|  | Cough/expectoration | 438 (87.8) | 630 (84.5) | 1068 (85.8) | 0.100 |
|  | Stuffy/running nose | 401 (80.4) | 584 (78.3) | 985 (79.1) | 0.377 |
|  | Dyspnea | 87 (17.4) | 114 (15.3) | 201 (16.1) | 0.312 |
|  | Fatigue | 349 (69.9) | 485 (65.0) | 834 (67.0) | 0.070 |
| COVID-19 vaccination | |  |  |  |  |
|  | Yes (n = 1208) | 478 (95.8) | 730 97.9) | 1208 (97.0) | 0.036 |
|  | No (n = 37) | 21 (4.2) | 16 (2.1) | 37 (3.0) |  |

^#^ Comorbidity or special condition includes hypertension, diabetes, cardiovascular disease, cerebrovascular disease, neoplastic disease, immune deficiency, chronic kidney disease, thyroid disease, rheumatoid arthritis, spinal joint disease, bronchial asthma, mental illness, and the third pregnancy and perinatal period

^*^ Multiple response

**S8 Association between** **parageusia and demographic/COVID-19-related characteristics among patients**

| Variables | | Parageusia (%) | | Total (%) | p value |
| --- | --- | --- | --- | --- | --- |
|  |  | Yes (n = 282) | No (n = 963) | N = 1245 |  |
| Sex | |  |  |  |  |
|  | Male | 50 (17.7) | 212 (22.0) | 262 (21.0) | 0.121 |
|  | Female | 232 (82.3) | 751 (78.0) | 983 (79.0) |  |
| Age, yr | |  |  |  |  |
|  | 18-29 | 223 (79.1) | 806 (83.7) | 1029 (82.7) | 0.006 |
|  | 30-39 | 48 (17.0) | 106 (11.0) | 154 (12.4) |  |
|  | 40-49 | 5 (1.8) | 41 (4.3) | 46 (3.7) |  |
|  | ≥50 | 6 (2.1) | 10 (1.0) | 16 (1.3) |  |
| Smoking | |  |  |  |  |
|  | Current | 14 (5.0) | 23 (2.4) | 37 (3.0) | 0.053 |
|  | Former | 13 (4.6) | 34 (3.5) | 47 (3.8) |  |
|  | Never | 255 (90.4) | 906 (94.1) | 1161 (93.3) |  |
| Alcohol | |  |  |  |  |
|  | Yes | 153 (54.3) | 472 (49.0) | 625 (50.2) | 0.122 |
|  | No | 129 (45.7) | 491 (51.0) | 620 (49.8) |  |
| Chronic rhinitis | |  |  |  |  |
|  | Yes | 39 (13.8) | 146 (15.2) | 185 (14.9) | 0.580 |
|  | No | 243 (86.2) | 817 (84.8) | 1060 (85.1) |  |
| Allergic rhinitis | |  |  |  |  |
|  | Yes | 78 (27.7) | 221 (22.9) | 299 (24.0) | 0.103 |
|  | No | 204 (72.3) | 742 (77.1) | 946 (76.0) |  |
| Any comorbidity or special condition^#^ | |  |  |  |  |
|  | Yes | 42 (14.9) | 106 (11.0) | 148 (11.9) | 0.076 |
|  | No | 240 (85.1) | 857 (89.0) | 1097 (88.1) |  |
| Method of diagnosis | |  |  |  |  |
|  | PCR test | 39 (13.8) | 162 (16.8) | 201 (16.1) | 0.484 |
|  | Antigen test | 154 (54.6) | 505 (52.4) | 659 (52.9) |  |
|  | Symptoms | 89 (31.6) | 296 (30.7) | 385 (30.9) |  |
| Rehabilitation status | |  |  |  |  |
|  | Complete | 98 (34.8) | 352 (36.6) | 450 (36.1) | 0.580 |
|  | Partial | 184 (65.2) | 611 (63.4) | 795 (63.9) |  |
| Symptoms^*^ | |  |  |  |  |
|  | Fever | 266 (94.3) | 910 (94.5) | 1176 (94.5) | 0.913 |
|  | Lack of appetite | 198 (70.2) | 517 (53.7) | 715 (57.4) | < 0.001 |
|  | Throat dryness and sore | 232 (82.3) | 744 (77.3) | 976 (78.4) | 0.072 |
|  | Myalgia | 211 (74.8) | 633 (65.7) | 844 (67.8) | 0.004 |
|  | Headache | 199 (70.6) | 655 (68.0) | 854 (68.6) | 0.417 |
|  | Diarrhea | 73 (25.9) | 189 (19.6) | 262 (21.0) | 0.023 |
|  | Cough/expectoration | 247 (87.6) | 821 (85.3) | 1068 (85.8) | 0.324 |
|  | Stuffy/running nose | 213 (75.5) | 772 (80.2) | 985 (79.1) | 0.092 |
|  | Dyspnea | 64 (22.7) | 137 (14.2) | 201 (16.1) | 0.001 |
|  | Fatigue | 208 (73.8) | 626 (65.0) | 834 (67.0) | 0.006 |
| COVID-19 vaccination | |  |  |  |  |
|  | Yes (n = 1208) | 276 (97.9) | 932 (96.8) | 1208 (97.0) | 0.342 |
|  | No (n = 37) | 6 (2.1) | 31 (3.2) | 37 (3.0) |  |

^#^ Comorbidity or special condition includes hypertension, diabetes, cardiovascular disease, cerebrovascular disease, neoplastic disease, immune deficiency, chronic kidney disease, thyroid disease, rheumatoid arthritis, spinal joint disease, bronchial asthma, mental illness, and the third pregnancy and perinatal period

^*^ Multiple response

**S9 Association between** **phantogeusia and demographic/COVID-19-related characteristics among patients**

| Variables | | Phantogeusia (%) | | Total (%) | p value |
| --- | --- | --- | --- | --- | --- |
|  |  | Yes (n = 333) | No (n = 912) | N = 1245 |  |
| Sex | |  |  |  |  |
|  | Male | 55 (16.5) | 207 (22.7) | 262 (21.0) | 0.018 |
|  | Female | 278 (83.5) | 705 (77.3) | 983 (79.0) |  |
| Age, yr | |  |  |  |  |
|  | 18-29 | 279 (83.8) | 750 (82.2) | 1029 (82.7) | 0.071 |
|  | 30-39 | 43 (12.9) | 111 (12.2) | 154 (12.4) |  |
|  | 40-49 | 5 (1.5) | 41 (4.5) | 46 (3.7) |  |
|  | ≥50 | 6 (1.8) | 10 (1.1) | 16 (1.3) |  |
| Smoking | |  |  |  |  |
|  | Current | 12 (3.6) | 25 (2.7) | 37 (3.0) | 0.364 |
|  | Former | 16 (4.8) | 31 (3.4) | 47 (3.8) |  |
|  | Never | 305 (91.6) | 856 (93.9) | 1161 (93.3) |  |
| Alcohol | |  |  |  |  |
|  | Yes | 170 (51.1) | 455 (49.9) | 625 (50.2) | 0.717 |
|  | No | 163 (48.9) | 457 (50.1) | 620 (49.8) |  |
| Chronic rhinitis | |  |  |  |  |
|  | Yes | 61 (18.3) | 124 (13.6) | 185 (14.9) | 0.038 |
|  | No | 272 (81.7) | 788 (86.4) | 1060 (85.1) |  |
| Allergic rhinitis | |  |  |  |  |
|  | Yes | 80 (24.0) | 219 (24.0) | 299 (24.0) | 0.997 |
|  | No | 253 (76.0) | 693 (76.0) | 946 (76.0) |  |
| Any comorbidity or special condition^#^ | |  |  |  |  |
|  | Yes | 40 (12.0) | 108 (11.8) | 148 (11.9) | 0.935 |
|  | No | 293 (88.0) | 804 (88.2) | 1097 (88.1) |  |
| Method of diagnosis | |  |  |  |  |
|  | PCR test | 50 (15.0) | 151 (16.6) | 201 (16.1) | 0.702 |
|  | Antigen test | 175 (52.6) | 484 (53.1) | 659 (52.9) |  |
|  | Symptoms | 108 (32.4) | 277 (30.4) | 385 (30.9) |  |
| Rehabilitation status | |  |  |  |  |
|  | Complete | 113 (33.9) | 337 (37.0) | 450 (36.1) | 0.327 |
|  | Partial | 220 (66.1) | 575 (63.0) | 795 (63.9) |  |
| Symptoms^*^ | |  |  |  |  |
|  | Fever | 317 (95.2) | 859 (94.2) | 1176 (94.5) | 0.492 |
|  | Lack of appetite | 231 (69.4) | 484 (53.1) | 715 (57.4) | < 0.001 |
|  | Throat dryness and sore | 264 (79.3) | 712 (78.1) | 976 (78.4) | 0.646 |
|  | Myalgia | 252 (75.7) | 592 (64.9) | 844 (67.8) | < 0.001 |
|  | Headache | 253 (76.0) | 601 (65.9) | 854 (68.6) | 0.001 |
|  | Diarrhea | 93 (27.9) | 169 (18.5) | 262 (21.0) | < 0.001 |
|  | Cough/expectoration | 294 (88.3) | 774 (84.9) | 1068 (85.8) | 0.126 |
|  | Stuffy/running nose | 269 (80.8) | 716 (78.5) | 985 (79.1) | 0.383 |
|  | Dyspnea | 72 (21.6) | 129 (14.1) | 201 (16.1) | 0.002 |
|  | Fatigue | 251 (75.4) | 583 (63.9) | 834 (67.0) | < 0.001 |
| COVID-19 vaccination | |  |  |  |  |
|  | Yes (n = 1208) | 321 (96.4) | 887 (97.3) | 1208 (97.0) | 0.428 |
|  | No (n = 37) | 12 (3.6) | 25 (2.7) | 37 (3.0) |  |

^#^ Comorbidity or special condition includes hypertension, diabetes, cardiovascular disease, cerebrovascular disease, neoplastic disease, immune deficiency, chronic kidney disease, thyroid disease, rheumatoid arthritis, spinal joint disease, bronchial asthma, mental illness, and the third pregnancy and perinatal period

^*^ Multiple response
